# Identifying Communities at Risk for Poor Health using Multidimensional vs. Unidimensional Neighborhood Disadvantage Indices

**DOI:** 10.64898/2026.08.06.26359856

**Authors:** Philippa Clarke, Kimberly Rollings, Robert Melendez, Kate Duchowny, Lindsay Gypin, Grace Noppert

**Affiliations:** Institute for Social Research, University of Michigan, Ann Arbor, MI, USA; Department of Epidemiology, School of Public Health, University of Michigan, Ann Arbor, MI, USA

**Keywords:** Neighborhood, socioeconomic status, disadvantage, deprivation, United States

## Abstract

**Background:** Neighborhood disadvantage indices used in public health research and policy include multiple economic, social, and housing items. However, research has failed to question whether it is necessary to include a multitude of economic, social, and housing variables in a single index. The purpose of this work was to examine three different neighborhood indices: a multidimensional disadvantage index, a unidimensional disadvantage index, and a unidimensional affluence index, and examine their performance with respect to distinguishing between healthy and unhealthy census tract neighborhoods in the United States.

**Methods:** The 2022 disadvantage and affluence indices came from the National Neighborhood Data Archive, which are derived from census tract data from the American Community Survey 5-year estimates (2018-2022). The multidimensional disadvantage index included seven economic, social (e.g., single parent households), and housing items; the unidimensional disadvantage index included three poverty and income items; the unidimensional affluence index included 3 items capturing greater social and economic resources. Data on neighborhood health status (census tract prevalence of obesity, diabetes, and coronary heart disease) was obtained from the Population Level Analysis and Community EStimates database for 2022 and linked to the disadvantage and affluence indices for 83,522 census tracts. Contingency tables examined the degree of correspondence in quintiles across the three different indices and the corresponding disease prevalence in each cell. Generalized linear mixed models regressed the disease prevalence variables on index quintiles to determine the predicted prevalence of disease across the disadvantage gradient for each index.

**Results:** Compared to the unidimensional disadvantage and affluence indices, the multidimensional disadvantage index underestimated disease burden in the most disadvantaged census tracts, and overestimated disease burden in the least disadvantaged tracts.

**Conclusions:** Using a disadvantage or affluence index with a more parsimonious set of items would have greater precision in identifying communities at risk for poor health.

## INTRODUCTION

Research has shown that residence in socioeconomically disadvantaged neighborhoods is associated with poor health.(1–4) Neighborhood disadvantage (also termed area-level deprivation or vulnerability) is hypothesized to shape health directly (e.g., through stress processes) (5,6) and indirectly (e.g., through a lack of resources to promote healthy behaviors, few opportunities for employment and education, and poor quality housing) (7–11). Area-level disadvantage is typically measured using administrative data on social, economic, and housing characteristics at local geographies (e.g., United States (US) census tract counts of people in poverty). While some research has relied on single indicator variables to capture disadvantage, composite measures that combine multiple social, economic, housing, and demographic variables in a single index have been shown to be more strongly associated with health outcomes (12,13). Composite indices capture the overlapping contexts that characterize local neighborhoods. Using a composite index in statistical analysis also avoids the risk of multicollinearity when using several related individual variables in the same model.

There are numerous composite disadvantage indices used in US clinical and public health research (13–15). These indices include multiple economic, social, and housing variables (ranging from 7-29 variables) at the local area level (e.g., census tract, block group), which are combined to create a single multidimensional index of disadvantage (13,16). For example, the Area Deprivation Index (ADI), one of the most widely used indices in clinical and health policy work, includes 17 census block group indicators capturing economic characteristics (family income, poverty), social characteristics (education, unemployment, single parent families) and housing characteristics (home ownership, home value, mortgage and rent, household crowding, plumbing, etc.) (17,18). Other commonly used indices, such as the Childhood Opportunity Index (COI)(19), the Social Vulnerability Index (SVI) (20), and the Social Deprivation Index (SDI) (21), capture similar economic, social, and housing items in their index construction in addition to other sociodemographic and environmental indicators (e.g., age structure, transportation, industrial pollutants). However, little research has questioned the necessity of including a multitude of economic, social, and housing variables in a neighborhood disadvantage index, or whether a more parsimonious index is sufficient to understand health outcomes.

A growing number of studies have systematically compared disadvantage indices to evaluate their ability to discriminate between good and poor health at the local area level (22–27). For example, Lou et al (2023) compared five US disadvantage indices on their relationships to 24 diverse health outcomes.(28) They found that area-level poverty, education, and employment were largely driving neighborhood effects on physical health, mental health, and mortality outcomes. Variables capturing neighborhood housing quality, demographic composition (language, minority status) and transportation (vehicle ownership) were less strongly associated with health, suggesting that including multiple social and environmental variables in disadvantage indices may be unnecessary, and could diminish their ability to identify local areas at risk for poor health.

Current health research on neighborhood disadvantage has also tended to overlook the concept of neighborhood affluence. Distinguished from non-disadvantaged neighborhoods by their large share of highly educated adults in professional occupations, affluent neighborhoods have the social and economic resources to promote and sustain local institutions that benefit the health of the neighborhood as a whole (29,30). This upper end of the neighborhood socioeconomic continuum, neighborhood affluence, has often been shown to be more important for health outcomes than neighborhood disadvantage (30–33). Yet, many disadvantage indices include indicators of affluence in their construction (e.g., ADI includes a variable for white collar occupations; COI includes variables for college education and high-skill employment), preventing the ability to distinguish the separate effects of disadvantage and affluence on health outcomes.

The purpose of this work was to examine three different neighborhood indices: a multidimensional disadvantage index (including economic, social, and housing variables), a unidimensional disadvantage index (including only economic variables), and a unidimensional affluence index, and examine their performance with respect to distinguishing between healthy and unhealthy census tract neighborhoods in the US. We evaluated the association between each of the three indices and the neighborhood prevalence of three costly and preventable adult health conditions (obesity, diabetes, and coronary heart disease) that have repeatedly been shown to be patterned by area-level disadvantage (8,34). We hypothesized that a multidimensional index would perform less well in identifying communities at risk for poor health compared to a unidimensional disadvantage or affluence index.

## METHODS

This is a national, cross-sectional study of US census tracts in 2022.

### Data and Measures

#### Neighborhood Disadvantage and Affluence

We used three indices from the National Neighborhood Data Archive (NaNDA) at the census tract level (35). Census tracts have a population size between 1,200 and 8,000 people, with an average size of 4,000 people, and are the preferred geographic level for studying the health effects of neighborhood disadvantage (14). Census tracts are also less likely to have missing data on income-related items compared to smaller geographic areas (i.e., block group), which are frequently suppressed by the Census Bureau, requiring imputation for indices created at the block group level. We used NaNDA’s 2022 disadvantage and affluence indices, which are derived from the US Census Bureau’s American Community Survey (ACS) 5-year estimates (2018–2022). These indices include census tract variables of economic, social, and housing characteristics that are combined into a mean index based on a principal components analysis and factor analysis conducted for each Census/ACS data release (36).

The <u>multidimensional disadvantage index</u> is the mean of seven variables: census tract proportions of people with annual income below the federal poverty level; proportion of households receiving public assistance income or food stamps; proportion of families with annual income <$40,000 (the lowest quartile of the national distribution of census tract family income in ACS 2018-2022); census tract proportion of people (age 16+) in the civilian labor force who are unemployed; proportion of people (age 25+) with less than high school degree; proportion of families with children that are single parents; and proportion of housing units that are not owner occupied. The <u>unidimensional disadvantage index</u> is the mean of three variables: census tract proportions of people with annual income below the federal poverty level; proportion of households receiving public assistance income or food stamps; and proportion of families with annual income <$40,000. The <u>unidimensional affluence index</u> is the mean of three variables: census tract proportions of people (age 25+) with a Bachelor’s Degree or higher; proportion of people (age 16+) in the civilian labor force who are employed in professional or managerial occupations (management, business, science, and arts occupations); and proportion of families with annual income >$125,000 (the highest quartile of the national distribution of census tract family income in ACS 2018-2022).

#### Neighborhood Health Status

Neighborhood health status was obtained from the CDC PLACES database for the year 2022 (37). The Population Level Analysis and Community EStimates (PLACES) database (38) includes crude prevalence of common health conditions for adults (age ≥18 years) nationwide at the census tract level since 2015. PLACES provides model-based health estimates using data from the Behavioral Risk Factor Surveillance System (BRFSS), Census decennial population counts and annual county population estimates, and the ACS 5-year estimates. Small-area estimation methods are used to obtain data on 12 health conditions and other health-related measures in US census tracts (37). The 2022 PLACES release includes data for 83,522 census tracts (at 2020 geography) in the 50 states and District of Columbia that have a Census 2020 adult population count ≥50. We used data on three adult health conditions: obesity ((BMI) ≥30.0 kg/m²), diabetes, and coronary heart disease (including angina). We used the census tract crude prevalence of these conditions (percent), rescaled to between 0 and 1 for analysis. CDC PLACES data were linked to NaNDA’s disadvantage and affluence indices by census tract Federal Information Processing Standards (FIPS) codes for all 83,522 census tracts at 2020 Census geography.

### Statistical Analyses

Neighborhood indices were categorized into quintiles for analysis. For comparability with the disadvantage indices, affluence was reversed so that the highest quintile represents the least affluent census tracts while the lowest quintile represents the most affluent census tracts. Descriptive statistics (means, standard deviation (SD)), were first used to summarize the prevalence of the three neighborhood health conditions across gradients (quintiles) of each of the three indices. Contingency tables examined the degree of correspondence in quintiles across the three different indices and the corresponding disease prevalence in each cell. Generalized linear mixed models with a logit link and a beta distribution were then used to regress the disease prevalence variables (modeled as proportions) on quintiles of each neighborhood index. A random effect for US state was included to account for unmeasured factors at the state level that could shape the prevalence of disease at the neighborhood level. Models adjusted for population density (persons per square mile) to account for census tract population and urbanicity. All analyses were conducted in SAS (Version 9.4) and RStudio (Version 4.4.2).

## RESULTS

### Univariate and Bivariate Results

Table 1 presents the mean prevalence of the three health conditions in US neighborhoods, overall and by quintiles of the three indices. Overall, the census tract prevalence of obesity, diabetes, and coronary heart disease was 34.4%, 12.4% and 7.0%, respectively (Table 1). There were clear gradients in the prevalence of each health condition across quintiles of the neighborhood indices, with the highest prevalence of disease in the most disadvantaged tracts (Q5) and the lowest prevalence in the least disadvantaged tracts (Q1). However, there were differences in disease prevalence between the highest (Q5) and lowest (Q1) quintiles depending on the disadvantage index used (Table 1). For example, the difference in the prevalence of coronary heart disease in neighborhoods at the highest and lowest quintiles of the unidimensional disadvantage index was 2.2% (8.1 – 5.9%), and 2.9% between neighborhoods in the highest and lowest quintiles of the reverse-affluence index (8.3 – 5.4%). However, there was only a 1% difference in neighborhood coronary heart disease prevalence between tracts at the highest and lowest quintiles of the multidimensional disadvantage index (7.5 – 6.5%). Similarly, the difference in the prevalence of diabetes between neighborhoods at the highest and lowest quintiles of the reverse-affluence index and the unidimensional disadvantage index was 7.4% and 6.9%, respectively, but only 5.5% between the highest and lowest quintiles of the multidimensional disadvantage index.

**Table 1.** Prevalence of Health Conditions by Three Neighborhood Indices: United States 2022 (N=83,522 census tracts)

|  | Census Tract Prevalence (%) Mean (SD) |  |  |
| --- | --- | --- | --- |
|  | Obesity | Diabetes | Coronary Heart Disease |
| Overall | 34.4 (7.2) | 12.4 (3.8) | 7.0 (2.2) |
| **Multidimensional Disadvantage Index** |  |  |  |
| Q1 (N=16,861) | 30.0 (5.2) | 10.3 (2.1) | 6.5 (1.8) |
| Q2 (N=16,886) | 32.5 (5.7) | 11.3 (2.4) | 7.0 (2.0) |
| Q3 (N=16,868) | 33.9 (6.2) | 12.0 (3.0) | 7.1 (2.2) |
| Q4 (N=16,814) | 35.7 (6.6) | 12.9 (3.6) | 7.1 (2.3) |
| Q5 (N=16,082) | 40.4 (7.4) | 15.8 (4.8) | 7.5 (2.4) |
| **Unidimensional Disadvantage Index** |  |  |  |
| Q1 (N=16,821) | 28.7 (5.3) | 9.6 (2.1) | 5.9 (1.7) |
| Q2 (N=16,856) | 31.6 (5.5) | 10.8 (2.3) | 6.6 (1.9) |
| Q3 (N=16,848) | 34.0 (5.4) | 12.0 (2.5) | 7.1 (2.0) |
| Q4 (N=16,839) | 36.5 (5.5) | 13.4 (2.9) | 7.6 (2.2) |
| Q5 (N=15,972) | 41.6 (6.6) | 16.5 (4.4) | 8.1 (2.3) |
| **Unidimensional Affluence Index (Reversed)** |  |  |  |
| Q1 (N=16,860) | 26.7 (4.7) | 9.1 (2.2) | 5.4 (1.7) |
| Q2 (N=16,828) | 31.5 (4.7) | 10.8 (2.2) | 6.4 (1.8) |
| Q3 (N=16,808) | 34.8 (4.8) | 12.2 (2.5) | 7.1 (1.9) |
| Q4 (N=16,641) | 37.7 (4.9) | 13.8 (2.9) | 7.9 (2.0) |
| Q5 (N=16,374) | 41.7 (5.8) | 16.5 (3.9) | 8.3 (2.2) |

Source: CDC PLACES data 2022 SD = standard deviation

In order to further quantify these differences, we examined the degree of correspondence in neighborhood quintiles across the three different indices. Table 2 presents contingency tables showing the number of census tracts where quintiles of the disadvantage and affluence indices agreed and where they disagreed in their classification. Supplementary Tables 1-3 present the corresponding disease prevalence (coronary heart disease, diabetes, and obesity, respectively) for each of these cross-classified quintiles. Consider, for example, the 16,907 census tracts classified as the most disadvantaged (Q5) by the multidimensional disadvantage index (Table 2a). The unidimensional disadvantage index also classified 82% (N=13,961) of these tracts as the most disadvantaged (Q5), and the average prevalence of coronary heart disease in these tracts was 7.9% (Supplemental Table 1a). However, in the remaining 18% of tracts that were considered less disadvantaged (Q1-Q4) by the unidimensional index but still the most disadvantaged (Q5) by the multidimensional index, the average prevalence of coronary heart disease was lower (5.7% on average, ranging from 3.9% to 5.8%, Supplemental Table 1a). Similarly, of the 16,907 census tracts classified as the most disadvantaged (Q5) by the multidimensional disadvantage index, only 61% (N=10,272) were classified as the most disadvantaged (Q5) by the reverse affluence index (Table 2b). In tracts where both the reverse affluence and multidimensional disadvantage indices agreed (both Q5), the average prevalence of coronary heart disease was 8.2% (Supplemental Table 1b). But in the remaining 6,635 tracts considered less disadvantaged (Q1-Q4) by the affluence index but highly disadvantaged (Q5) by the multidimensional index, the average prevalence of coronary heart disease was lower at 6.5% (ranging from 4.1% to 7.2%, Supplemental Table 1b). Thus, the multidimensional disadvantage index classified about 40% of US census tracts as highly disadvantaged when in fact they were less disadvantaged in terms of their health status.

**Table 2a.** Contingency Table of US Census Tracts by Quintiles of Multidimensional vs. Unidimensional Disadvantage Index: ACS 2018-22, N=84,539.

|  | Unidimensional Disadvantage Index Quintiles |  |  |  |  |  |  |
| --- | --- | --- | --- | --- | --- | --- | --- |
| Multidimensional Disadvantage Index Quintiles | Q1 | Q2 | Q3 | Q4 | Q5 | Missing | Total |
| Q1 | 12435 | 4192 | 208 | 0 | 0 | 72 | 16907 |
| Q2 | 3254 | 8167 | 5181 | 296 | 1 | 9 | 16908 |
| Q3 | 967 | 3656 | 7525 | 4681 | 66 | 12 | 16907 |
| Q4 | 199 | 842 | 3738 | 9256 | 2840 | 33 | 16908 |
| Q5 | 13 | 11 | 216 | 2635 | 13961 | 71 | 16907 |
| Missing |  |  |  |  |  | 2 | 2 |
| Total | 16868 | 16868 | 16868 | 16868 | 16868 | 199 | 84539 |

**Table 2b.** Contingency Table of US Census Tracts by Quintiles of Multidimensional Disadvantage Index and Unidimensional Affluence (Reversed): ACS 2018-22 (N=84,539)

|  | Unidimensional Affluence Index (Reversed) Quintiles |  |  |  |  |  |  |
| --- | --- | --- | --- | --- | --- | --- | --- |
| Multidimensional Disadvantage Index Quintiles | Q1 | Q2 | Q3 | Q4 | Q5 | Missing | Total |
| Q1 | 8964 | 5259 | 2110 | 482 | 92 |  | 16907 |
| Q2 | 3753 | 5092 | 4798 | 2822 | 443 |  | 16908 |
| Q3 | 2477 | 3441 | 4500 | 4717 | 1772 |  | 16907 |
| Q4 | 1454 | 2237 | 3736 | 5153 | 4328 |  | 16908 |
| Q5 | 259 | 878 | 1764 | 3733 | 10272 | 1 | 16907 |
| Missing |  |  |  |  |  | 2 | 2 |
| Total | 16907 | 16907 | 16908 | 16907 | 16907 | 3 | 84539 |

**Table 2c.** Contingency Table of US Census Tracts by Quintiles of Unidimensional Disadvantage Index and Unidimensional Affluence (Reversed): ACS 2018-22 (N=84,539)

|  | Unidimensional Affluence Index (Reversed) Quintiles |  |  |  |  |  |  |
| --- | --- | --- | --- | --- | --- | --- | --- |
| Unidimensional Disadvantage Index Quintiles | Q1 | Q2 | Q3 | Q4 | Q5 | Missing | Total |
| Q1 | 10539 | 4704 | 1302 | 260 | 63 |  | 16868 |
| Q2 | 4140 | 6153 | 4610 | 1709 | 256 |  | 16868 |
| Q3 | 1486 | 3682 | 5633 | 4745 | 1322 |  | 16868 |
| Q4 | 570 | 1739 | 3815 | 6239 | 4505 |  | 16868 |
| Q5 | 159 | 611 | 1538 | 3944 | 10615 | 1 | 16868 |
| Missing | 13 | 18 | 10 | 10 | 146 | 2 | 199 |
| Total | 16907 | 16907 | 16908 | 16907 | 16907 | 3 | 84539 |

At the other end of the disadvantage spectrum, of the 16,907 census tracts classified as the least disadvantaged (lowest quintile Q1) according to the multidimensional disadvantage index, just over half of these tracts (N=8,964) were also classified as the least disadvantaged (Q1) by the reverse affluence index (Table 2b). The average prevalence of coronary heart disease in these tracts was 5.9% (Supplemental Table 1b). But in the remaining 7,943 census tracts classified as the least disadvantaged (Q1) by the multidimensional index (but more disadvantaged [Q2-Q5] by the affluence index), coronary heart disease prevalence was higher (ranging from 6.7% to 8.1%, Supplemental Table 1b). There was greater agreement in the classification of the least disadvantaged tracts (Q1) between the multidimensional disadvantage and unidimensional disadvantage indices (74% of the tracts classified as the least disadvantaged by the multidimensional index were also classified as the least disadvantaged by the unidimensional index, Table 2a), where the mean prevalence of coronary heart disease was 6.1% (Supplemental Table 1a). However, in the remaining 4,400 census tracts classified as the least disadvantaged by the multidimensional index but more disadvantaged (Q2-Q3) by the unidimensional index, the prevalence of disease was much higher (ranging from 7.4% to 9.1%, Supplemental Table 1a), suggesting that these neighborhoods are in fact more disadvantaged in terms of health than the multidimensional index would indicate.

### Regression Results

Results from the generalized linear mixed models regressing each health outcome on each of the three disadvantage indices are presented in Supplemental Table 4. For ease of interpretation, Figures 1-3 present the predicted prevalence from these models (population average census tract prevalence with 95% CI) for obesity, diabetes, and coronary heart disease (respectively) at average population density across quintiles of each of the disadvantage indices (again, affluence is reverse-scored). Overall, across all three indices, the prevalence of obesity, diabetes, and coronary heart disease was highest at greater levels of disadvantage and lowest at lower levels of disadvantage. However, there were differences in the predicted prevalence of disease at each disadvantage quintile depending on the index used. For example (Figure 1), in the least disadvantaged tracts as measured by the multidimensional disadvantage index, there was 2.3% higher prevalence of obesity than in the least disadvantaged tracts as measured by the reverse affluence index (quintile 1 multidimensional disadvantage index = 29.7% (95% CI: 28.7, 30.6) vs. quintile 1 reverse affluence index = 27.4% (95% CI=26.7, 28.2)). Similarly, the prevalence of diabetes was 10% (95% CI=9.5, 10.4) in the least disadvantaged tracts when using the multidimensional disadvantage index, but only 8.8% (95% CI=8.5, 9.2) in the least disadvantaged tracts when using the affluence index (reversed) (Figure 2). For coronary heart disease (Figure 3), the neighborhood prevalence was 6.4% (95% CI=6.2, 6.6) in the least disadvantaged tracts as measured by the multidimensional disadvantage index compared to 5.9% (95% CI=5.7, 6.1) when using the unidimensional disadvantage index, and 5.6% (95% CI=5.5, 5.8) in the least disadvantaged tracts when using the affluence index (reversed). In general, the prevalence of disease in the least disadvantaged tracts tended to be lowest when using the affluence index and highest when using the multidimensional disadvantage index.

**Figure 1.**
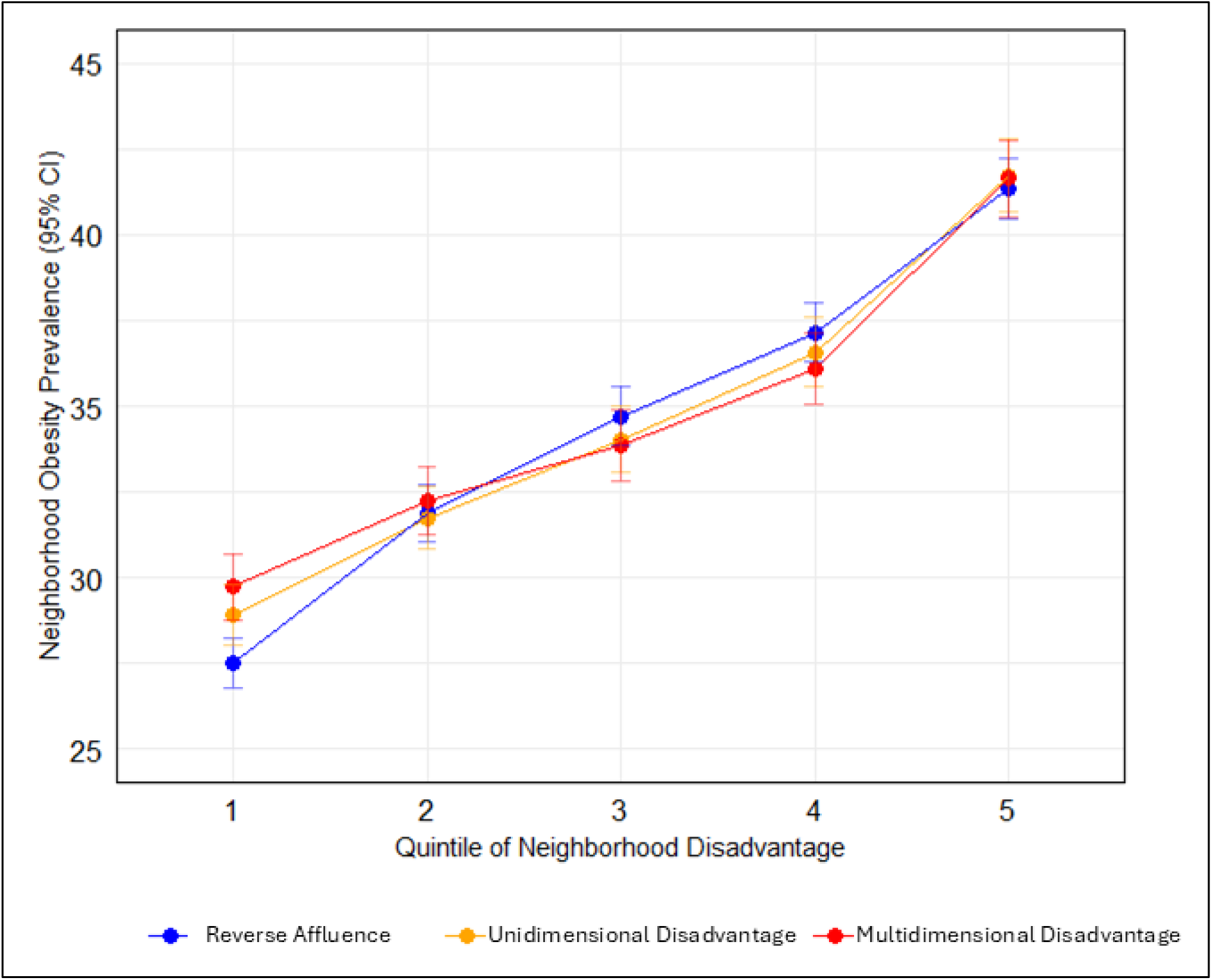
Predicted Census Tract Prevalence (%) of Obesity by Quintiles of Neighborhood Disadvantage Indices (United States, 2022) Note: predicted prevalence is population average effect adjusted for census tract population density. All models include a random effect State Source: CDC PLACES data 2022

**Figure 2.**
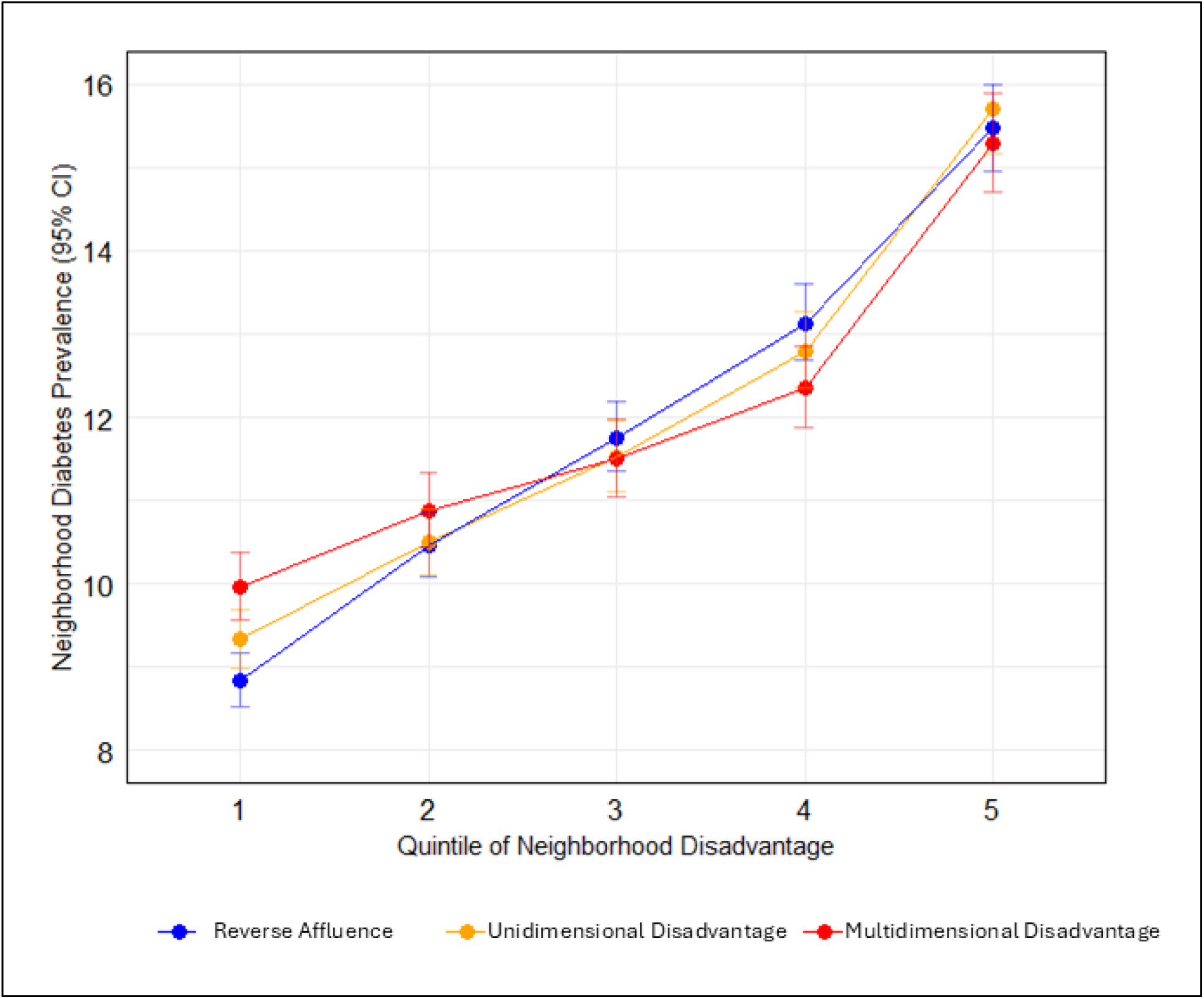
Predicted Census Tract Prevalence (%) of Diabetes by Quintiles of Neighborhood Disadvantage Indices (United States, 2022) Note: predicted prevalence is population average effect adjusted for census tract population density. All models include a random effect State Source: CDC PLACES data 2022

**Figure 3.**
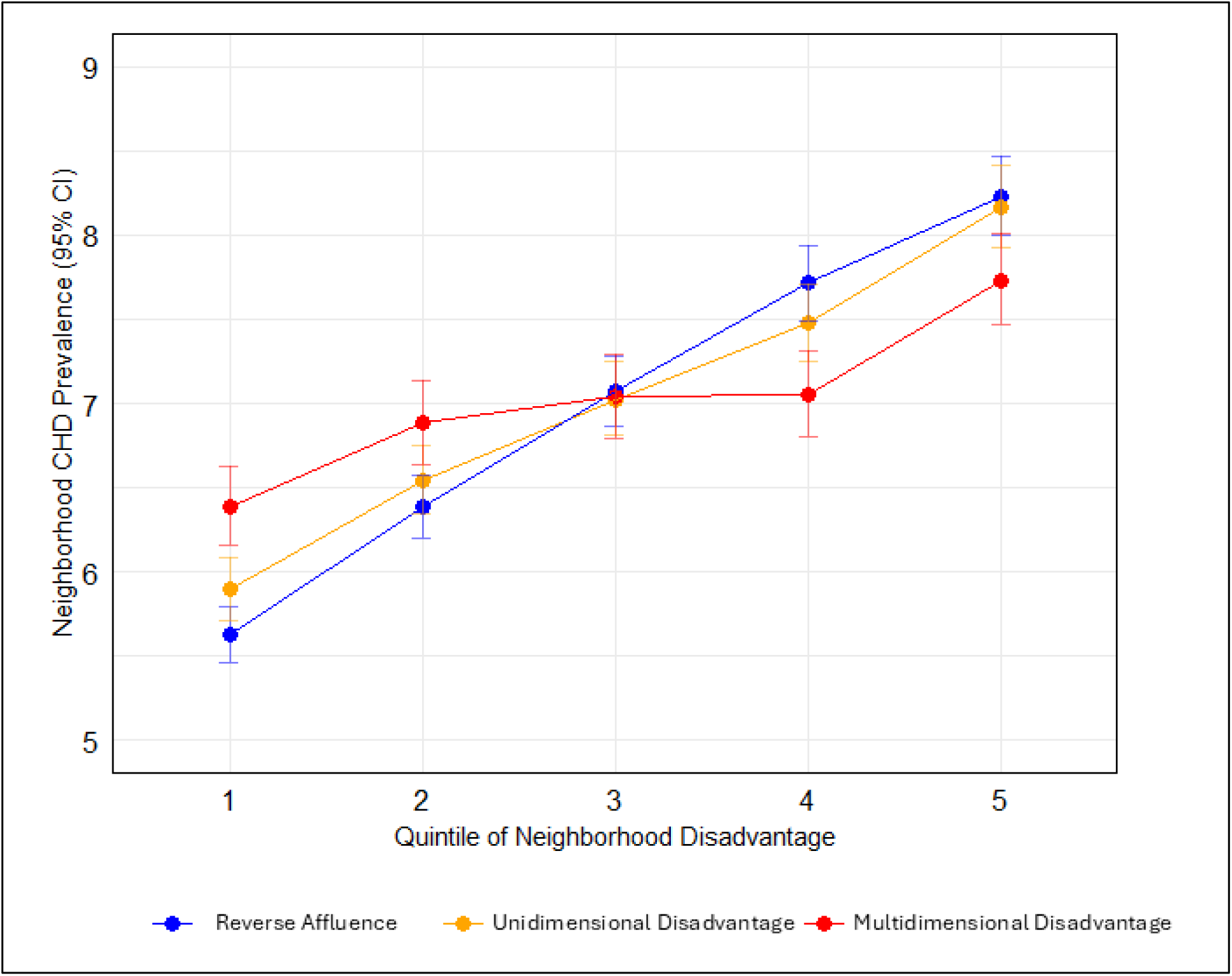
Predicted Census Tract Prevalence (%) of Coronary Heart Disease (CHD) by Quintiles of Neighborhood Disadvantage Indices (United States, 2022) Note: predicted prevalence is population average effect adjusted for census tract population density. All models include a random effect State Source: CDC PLACES data 2022

At higher levels of neighborhood disadvantage, there was a convergence in the predicted prevalence of disease around the middle quintiles for all three indices. But by the third quintile of disadvantage, the predicted prevalence of disease was lower in tracts classified by the multidimensional disadvantage index compared to the unidimensional disadvantage index and affluence index. For example (Figure 3), at the fourth quintile of disadvantage the predicted prevalence of coronary heart disease was 7.1% (95% CI=6.8, 7.3) when using the multidimensional disadvantage index but was 0.6 percent higher (7.7%; 95% CI=7.5, 7.9) in census tracts classified in the fourth quintile of the reverse affluence index. The pattern was similar for the other conditions (diabetes and obesity), but the confidence intervals overlapped at higher levels of disadvantage. The unidimensional disadvantage index tended to track most closely with the reverse affluence index for the predicted prevalence of diabetes (Figure 2) and coronary heart disease (Figure 3), although the confidence intervals overlapped with both the affluence index and multidimensional disadvantage index. In general, the multidimensional disadvantage index tended to underestimate disease burden in the most disadvantaged census tracts and overestimate disease burden in the least disadvantaged tracts. The unidimensional disadvantage index and the affluence index provided greater precision in identifying the most disadvantaged and least disadvantaged neighborhoods with respect to health status.

## DISCUSSION

Using national data on the neighborhood prevalence of three chronic health conditions in the US, we compared disease prevalence across quintiles of three different disadvantage indices: a multidimensional disadvantage index (including economic, social, and housing variables), a unidimensional disadvantage index (based on indicators of poverty and low income), and a unidimensional index of neighborhood affluence (based on indicators of high income, education, and occupational position). We found that population health at the neighborhood level was patterned differently depending on the index used. In comparison to the multidimensional disadvantage index, the unidimensional affluence and disadvantage indices were better able to differentiate disease prevalence across the gradients in disadvantage.

There are at least 33 publicly available disadvantage indices in the US, which include multiple economic, social, and housing items (up to 29 items in a single index). (13–15) None of these indices distinguish between disadvantage and affluence. Yet, our results provided evidence that disadvantage may be best captured using a more parsimonious set of items with respect to identifying populations at risk for poor health. Compared to the unidimensional disadvantage or affluence indices (each with three variables), the multidimensional disadvantage index (with seven variables) had less precision in distinguish between neighborhoods with highest and lowest disease burden.

Findings have implications for the use of disadvantage indices in public health policy to identify populations most in need of resources. Neighborhood disadvantage indices have recently been used in US health policy to inform vaccine distribution (39) and to account for social risks in Medicare and Medicaid payments (15,40). For example, the Centers for Medicare and Medicaid Services (CMS) Accountable Care Organization Realizing Equity, Access, and Community Health (ACO REACH) program uses the Community Deprivation Index (CDI) to identify disadvantaged areas and provide additional resources to health care providers that serve these areas (41). The CDI is a multidimensional index with 17 variables capturing economic, education, family, household, transportation, and housing domains. However, our findings indicate that using a multidimensional index risks redirecting resources away from communities classified as the least disadvantaged that in fact have higher prevalence of disease in comparison to the least disadvantaged communities as defined by a unidimensional index. For example, in comparison to the unidimensional disadvantage index, using the multidimensional disadvantage index to identify low risk communities would direct resources away from over 4000 census tracts (Table 2a) representing over 16 million people (based on an average tract population of 4000) with higher disease prevalence of coronary heart disease (Supplemental Table 1a). Similarly, using a multidimensional disadvantage index instead of a unidimensional affluence index to identify communities in greatest need of health care would potentially misdirect resources towards over 6000 US census tract neighborhoods (Table 2b) representing over 24 million people, where coronary heart disease prevalence is in fact lower (Supplemental Table 1b). Using a unidimensional disadvantage or affluence index would result in greater precision in identifying communities in greatest need of health care resources.

### Strengths and Limitations

Strengths of this study include the nationwide analysis of all residential census tracts in the United States, and the rigorous evaluation of multidimensional vs. unidimensional indices as they relate to national estimates of neighborhood health conditions in 2022. Despite these strengths, some limitations should be acknowledged. The prevalence of neighborhood health conditions came from the CDC PLACES data, which are model-based estimates using small-area estimation methods (multilevel regression and post stratification) (37). While direct population-based survey estimates would have been ideal for our validity analyses, PLACES provides the only publicly available census tract data on chronic disease prevalence. Moreover, the PLACES model-based estimates have been shown to be strongly correlated with direct survey estimates from both BRFSS data and local survey data at the state, county, and city levels (42–45), supporting the validity of the data. Although the three disadvantage indices used in this study were empirically derived based on a factor analysis, (36) they were operationalized as a mean of all index items. Greater precision might be attained by using factor scores in index construction but could also detract from their ease of interpretation in public health policy and research.

## CONCLUSIONS

The inclusion of multiple social, economic, education, housing, and transportation items in area-based disadvantage indices may be compromising their performance with respect to understanding population health. Our findings suggest that a unidimensional affluence or disadvantage index with a more parsimonious set of items would have greater precision in identifying communities at risk for poor health in the US.

## DECLARATIONS

### Availability of data and materials

The data for the disadvantage indices is available at the Inter-University Consortium for Political and Social Research via https://doi.org/10.3886/ICPSR38528.v6

The CDC PLACES data are available at the following URL: https://data.cdc.gov/500-Cities-Places/PLACES-Local-Data-for-Better-Health-County-Data-20/swc5-untb/about_data

## Data Availability

The data are publicly available as follows: Data for the disadvantage indices is available from the Inter-university Consortium for Political and Social Research (ICPSR) via https://doi.org/10.3886/ICPSR38528.v5 The CDC PLACES data are available at the following URL: https://data.cdc.gov/500-Cities-Places/PLACES-Local-Data-for-Better-Health-County-Data-20/swc5-untb/about_data

https://www.icpsr.umich.edu/web/ICPSR/studies/38528/versions/V5

